# A corpus of GA4GH Phenopackets: case-level phenotyping for genomic diagnostics and discovery

**DOI:** 10.1101/2024.05.29.24308104

**Authors:** Daniel Danis, Michael J Bamshad, Yasemin Bridges, Pilar Cacheiro, Leigh C Carmody, Jessica X Chong, Ben Coleman, Raymond Dalgleish, Peter J Freeman, Adam S L Graefe, Tudor Groza, Julius O B Jacobsen, Adam Klocperk, Maaike Kusters, Markus S Ladewig, Anthony J Marcello, Teresa Mattina, Christopher J Mungall, Monica C Munoz-Torres, Justin T Reese, Filip Rehburg, Bárbara C S Reis, Catharina Schuetz, Damian Smedley, Timmy Strauss, Jagadish Chandrabose Sundaramurthi, Sylvia Thun, Kyran Wissink, John F Wagstaff, David Zocche, Melissa A Haendel, Peter N Robinson

## Abstract

The Global Alliance for Genomics and Health (GA4GH) Phenopacket Schema was released in 2022 and approved by ISO as a standard for sharing clinical and genomic information about an individual, including phenotypic descriptions, numerical measurements, genetic information, diagnoses, and treatments. A phenopacket can be used as an input file for software that supports phenotype-driven genomic diagnostics and for algorithms that facilitate patient classification and stratification for identifying new diseases and treatments. There has been a great need for a collection of phenopackets to test software pipelines and algorithms. Here, we present phenopacket-store. Version 0.1.12 of phenopacket-store includes 4916 phenopackets representing 277 Mendelian and chromosomal diseases associated with 236 genes, and 2872 unique pathogenic alleles curated from 605 different publications. This represents the first large-scale collection of case-level, standardized phenotypic information derived from case reports in the literature with detailed descriptions of the clinical data and will be useful for many purposes, including the development and testing of software for prioritizing genes and diseases in diagnostic genomics, machine learning analysis of clinical phenotype data, patient stratification, and genotype-phenotype correlations. This corpus also provides best-practice examples for curating literature-derived data using the GA4GH Phenopacket Schema.

## Main text

Over 10,000 rare diseases have been identified to date,^1^ collectively affecting between 3.5% and 8% of the population,^2^ yet many patients experience a long diagnostic odyssey of 5-7 years.^1,3^ Previously, each of the numerous software packages that support phenotype-driven genomic diagnostic have used bespoke input formats for phenotypic data, and information about the pedigree. The Phenopacket Schema provides a standard input format for such tools that will simplify computational analysis pipelines.

Ontologies are systematic representations of knowledge that can be used to capture medical phenotype data by providing concepts (terms) from a knowledge domain and additionally specifying formal semantic relations between the concepts. Ontologies enable precise patient classification by supporting the integration and analysis of large amounts of heterogeneous data.^4^ The Human Phenotype Ontology (HPO), developed by the Monarch Initiative,^5^ is widely used in human genetics and other fields that care for individuals with rare diseases,^6^ and is also increasingly being used in other settings such as Electronic Healthcare Records (EHRs).^7,8^ HPO terms represent phenotypic features such as signs, symptoms, as well as laboratory and imaging findings. However, the HPO itself does not specify how HPO terms and data should be arranged to record and exchange such information along with genomic data. To address this, in the context of the Global Alliance for Genomics and Health (GA4GH), we developed the Phenopacket Schema, a standard for sharing disease and phenotype information. A phenopacket is a computational representation of an individual person or biosample, linking that individual to phenotypic descriptions, genetic information, diagnoses, and treatments.^9,10^

The Phenopacket Schema allows clinical data (phenotypic attributes, measurements, treatments and other medical actions) from individual patients to be compared and shared broadly, in contrast to the sensitive clinical data found within EHRs and other contexts. Such comparisons can aid in diagnosis and facilitate patient classification and stratification for identifying new diseases and treatments.^11^ The Phenopacket Schema is designed to support interoperability between people, organizations, and systems to advance the worldwide effort to address human disease and biological understanding. These partners include clinical laboratories, authors, journals, clinicians, data repositories, patient registries, electronic health record (EHR) systems, and knowledge bases. The Phenopacket Schema does not model -omics data in detail but does enable users to link a phenopacket to files representing data from high-throughput screening techniques or to denote individual variants in several formats.^11^ The Phenopacket Schema integrates a version of the GA4GH Variant Representation Specification and is designed to be interoperable with other GA4GH standards, including those for pedigree data.^12^

The Phenopacket Schema aims to represent data from different sources, including data from EHRs, research studies, data entry tools, or published case reports, in a consistent and computable format to enable the sharing and integration of structured clinical data. The core principles of the schema include composability, traceability (data provenance), the FAIR principles (Findable, Accessible, Interoperable, and Reusable), and computability.^13^ Multiple upstream data collection and management tools already support exporting patient profiles as Phenopackets for downstream analysis and data sharing, including PhenoTips,^14^ RD-Connect Genome-Phenome Analysis Platform,^15^ Patient Archive in Australia, and IRUD Exchange in Japan.^16^ PhenoTips can generate Phenopackets from patient or family records through a user interface or REST APIs, and includes de-identified demographic data, clinical phenotype, diagnoses, curated genetic findings, and pedigree data.^17^ Exomiser,^18,19^ LIRICAL.^20^ SvAnna,^21^ Phen2Gene,^22^ and CADA^23^ already accept phenotype data in Phenopacket format. Projects such as EU-funded Solve-RD and the European Joint Programme on Rare Diseases (EJP-RD) can generate Phenopackets for the data included in GPAP, which aims to facilitate diagnosis and novel gene discovery for clinical researchers.^24^ Phenopackets are used in Solve-RD to share phenotypic and other relevant clinical or genetic information (e.g., candidate or causative variants) between the consortium members, and are also deposited along the genomics data at the European Genome-phenome Archive (EGA) for long-term archival and controlled access. Besides being a successful instrument for data import/export between the project’s databases, Phenopackets have also proved to be useful for data analysis, such as clustering patients based on their phenotypic similarity.^25^

There is a need for a collection of phenopackets to test the software pipelines and algorithms that work on individual rare and genetic disease patient cases. In this work, we have created phenopacket-store, a collection of 4916 Phenopackets with clinical data from individuals with one of 277 Mendelian and chromosomal diseases. We developed pyphetools, a Python package with functionality to streamline the creation of phenopackets from tabular data often found in the medical literature. We selected publications for curation from the human genetics literature to represent a broad range of diseases. Publications were considered if they present individual-level data about one or more individuals affected by a given disease. Publications were not included if they provided only aggregate or summary-level information. For instance, if 7/12 patients in some cohorts are reported to have *scoliosis* and 3/12 to have *pes planus*, but no information was provided about the specific features that each of the individuals in the cohort had, then the publication would not be a candidate for inclusion in phenopacket-store. A typical table contains information about patients in rows and one column for each data item (age of onset, sex, genetic variants, phenotypic features, etc.). For publications that do not contain such tables, pyphetools offers various helper functions that assist with manual curation and filling of an Excel template from which Phenopackets can then be created. The Phenopacket Schema is a model that can be stored in many formats. We recommend JSON and have stored each phenopacket in this repository as a JSON file.

One of the goals of the phenopacket-store project is to provide a collection of best-practice Phenopackets for rare genetic disease that will enable software developers to test program code and develop novel algorithms. We have curated a wide range of rare diseases including cohorts ranging from 1 to 463 individuals (Figure 1). Version 0.1.12 of Phenopacket-store comprises Phenopackets representing 4916 individuals with 277 diseases associated with variants in 236 genes. 2872 distinct variants are included, and the information was derived from 605 different publications. A total of 2732 distinct HPO terms were used. An average of 17.7 individuals were curated per disease (median 7). 16 genes in the collection were associated with two Mendelian diseases, and 8 genes were associated with more than two diseases.

**Figure 1.**
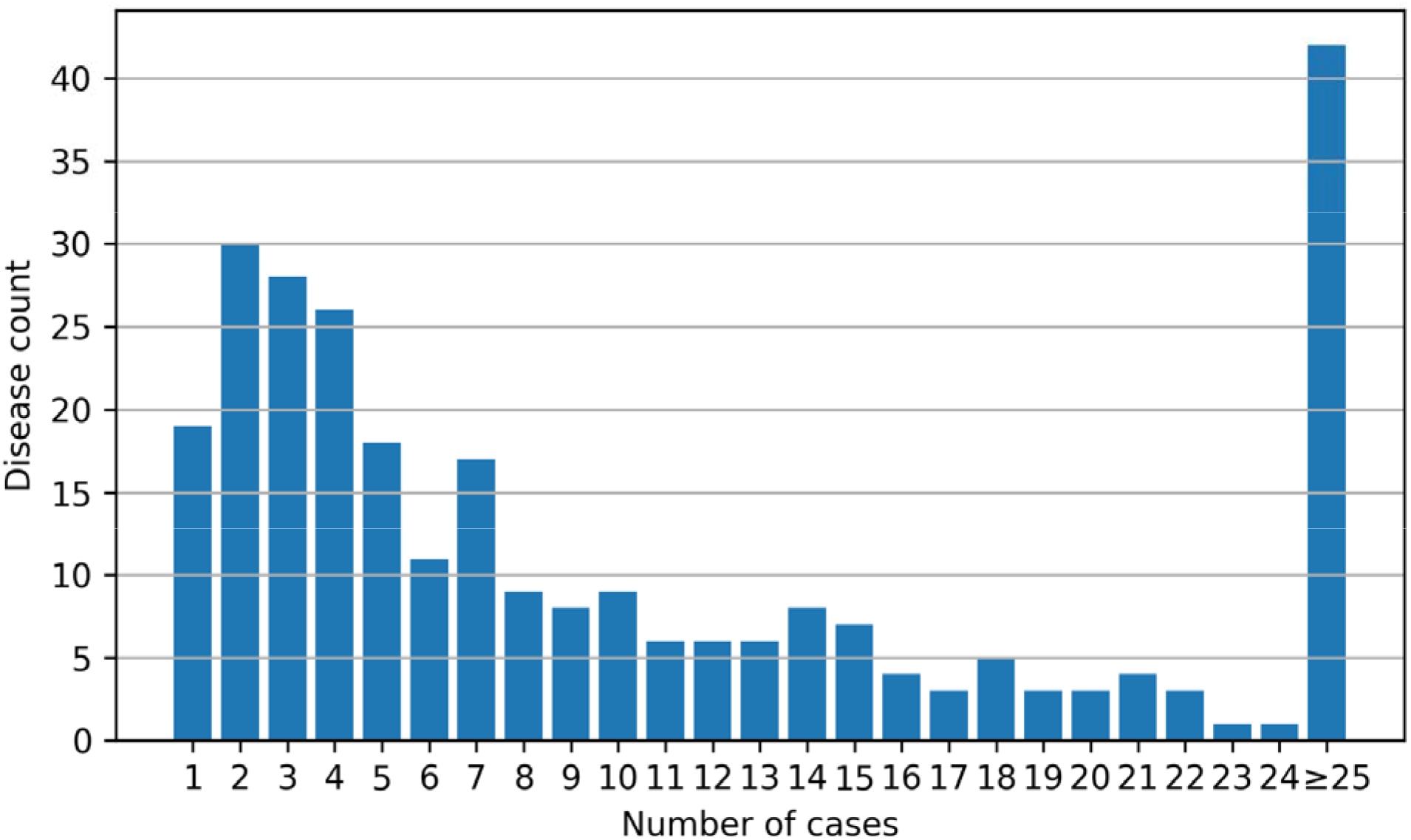
The plot shows the number of diseases for which the indicated number of cases is available (phenopacket-store, version 0.1.12).

The pyphetools library contains extensive quality-control code to prevent format errors. We additionally validate each of the Phenopackets using the Java command-line application called phenopacket-tools.^11^ We have created the Phenopackets with the following rules and assumptions:

### Phenopacket and individual identifiers

In the GA4GH Phenopacket Schema, both the phenopacket and the individual (patient) have identifiers. We have used the identifiers in the original publications for the individual id. If no identifier was provided, we used the word “individual”. Note that the individual id must be distinct for all individuals described in any publication. For the phenopacket id, we prepended the PubMed identifier. For instance, in a publication about variants in the VRK1 gene,^26^ an individual with the identifier BAB3022 was described. We use this for the individual id, and for the phenopacket id, we use PMID_24126608_BAB3022. The Phenopacket Schema does not require PubMed identifiers, but for this repository, we are only including published care reports with a PubMed identifier. In some cases, a single individual has been published several times with different identifiers (see, for instance, individual #00318253 in the Leiden Open Variation Database ^27^). It is outside the scope of the Phenopacket Schema to address the issue of duplication, but we recommend that curators be aware of this potential problem and take measures not to create multiple phenopackets that represent the same individual.

### Age of onset and age at last examination

Wherever possible, the age of onset was curated from the original publication (i.e., the age of the first manifestation of the disease). Additionally, the age at the latest examination was curated. Some Phenopackets additionally have information about the age of death (if applicable).

### Disease diagnosis

We encode the disease diagnosis in the top-level list of disease elements. The Phenopacket Schema does not specify which disease terminology should be used; use of the Online Mendelian Inheritance in Man (OMIM) identifiers^28^ or Mondo disease ontology identifiers are recommended.^29^ The age of onset, the age of manifestation of the first sign or symptom of a disease, is encoded as a part of the disease element. Because the current collection of Phenopackets is focused on representing published case reports with genetic diagnoses, the disease is also recorded as a part of the genomic interpretation element. The disease recorded here must match one of the diseases in the top-level disease list or an error will be recorded. Note that for other purposes, the top-level list of disease elements could record additional diseases or could use a Mondo term such as nonsyndromic genetic hearing loss (MONDO:0019497) to represent the clinical diagnosis made before genetic testing. We have not provided these candidate diagnoses in this collection of Phenopackets because, in general, the information is not available in the published clinical case reports. Figure 2 provides a simplified overview of the internal structure of a single phenopacket entry.

**Figure 2.**
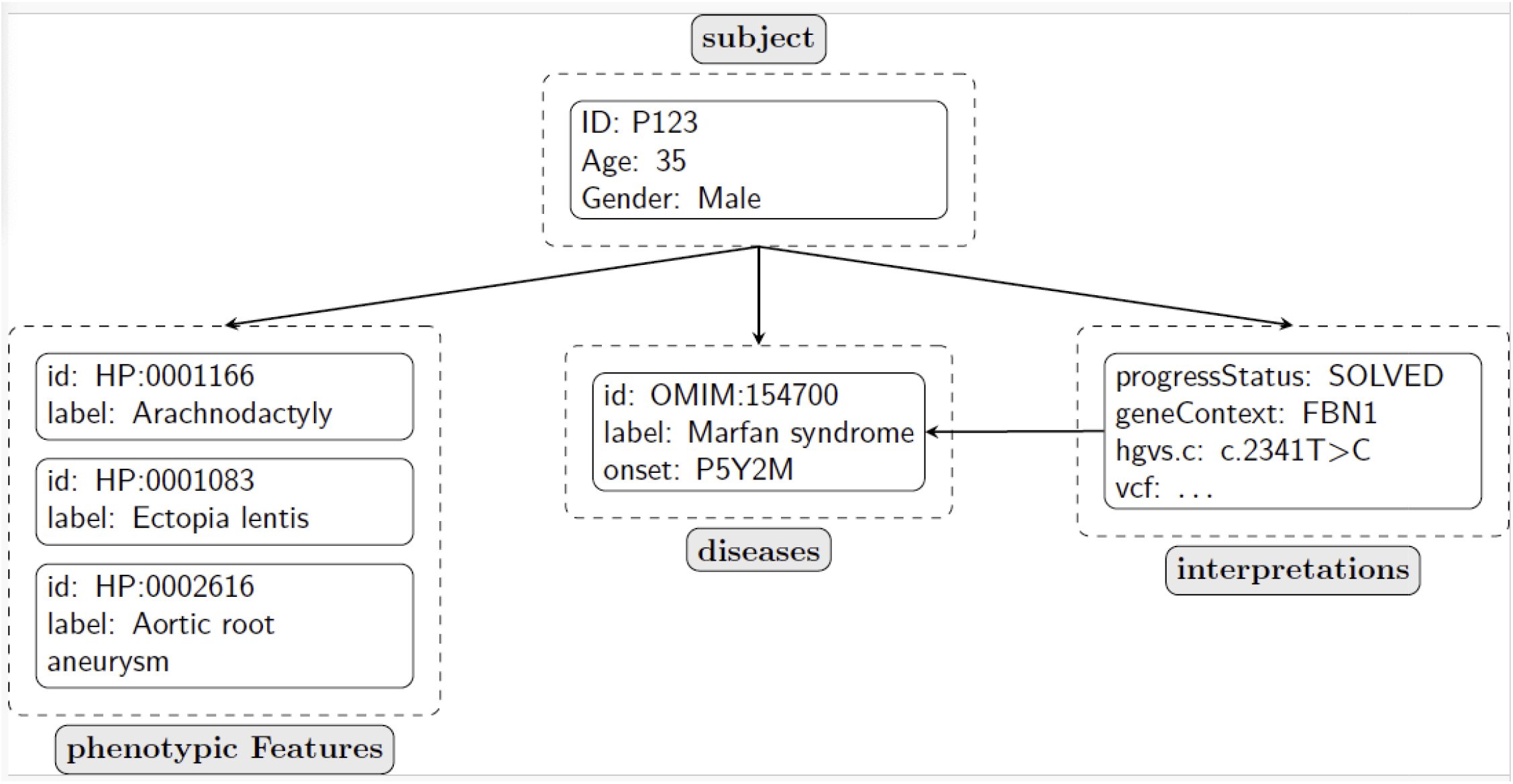
Schematic visualization of a phenopacket. In this simplified representation, the major elements of the Phenopacket Schema used for the Phenopackets in this collection are shown. The subject of the phenopacket is represented using the Individual element, which allows the (anonymous) identifier, age at last examination, and sex to be specified. Each subject can have an arbitrary number of phenotypic features, which comprise an HPO term and optionally information about the age of onset of the feature. The subject can have an arbitrary number of diseases, but for the phenopackets contained in this collection, each subject has one disease. The subject can have an arbitrary number of interpretations, which must refer to a disease in the disease list. In this example, a pathogenic variant in the *FBN1* gene is interpreted to be causal for Marfan syndrome.

Increasing the volume of computable data across a diversity of systems will support global computational disease analysis by integrating genotype, phenotype, and other multi-modal data for precision health applications. GA4GH Phenopackets can be generated from a variety of source data and can be used for many different kinds of analysis. Phenopackets intend to make data “analysis-ready” or “AI-ready,” so that software tools can perform various analytics tasks or queries across collections of phenopackets without extensive data transformations prior to the computational logic.

The Phenopacket Schema was designed to support a number of use cases in a range of fields including rare disease diagnostics, biobanking, oncology, and EHR integration. Here, we have created a substantial collection of phenopackets representing individuals diagnosed with a rare genetic disease. The collection is intended to be used by bioinformaticians and other analysts to develop and test software; for instance, the performance of a genomic diagnostic software could be tested by simulating cases using the phenopackets by spiking the causal variants reported in the phenopackets into VCF files that are representative of the population being tested. The collection also provides examples of best practices in creating phenopackets for databases, or to accompany manuscripts describing case reports or cohorts of individuals with a rare disease. Additionally, the Monarch Initiative is currently updating its HPO annotation pipeline to use phenopackets in addition to the previous HPO annotations file (phenotype.hpoa).^6^

The phenopackets in this repository represent the first large-scale collection derived from case reports in the literature with detailed descriptions of the clinical data. They will be useful for many purposes, including the development and testing of software for prioritizing genes and diseases in diagnostic genomics, machine learning analysis of clinical phenotype data, patient stratification, and genotype-phenotype correlations. They also provide a set of best-practices examples for curating literature-derived data using the GA4GH Phenopacket Schema. Genomic data will become ever more important in translational research and clinical care in the coming years and decades. The Phenopacket Schema represents a standard for capturing clinical data and integrating it with genomic data that will help to obtain the maximal utility of this data for understanding disease and developing precision medicine approaches to therapy.

## Data and code availability

Phenopacket-store is available at https://github.com/monarch-initiative/phenopacket-store under a BSD3 open-source license. The phenopackets generated with the phenopacket-store code are available under the “Releases” tab of the repository. Version 0.1.12 was presented in this manuscript.

Phenopacket-store makes use of the pyphetools library to create phenopackets. Pyphetools ios a Python library and is available at https://github.com/monarch-initiative/pyphetools under a MIT License. Version 0.9.85 was current at the time of this writing. Pyphetools is additionally available at the Python Package Index (pypi) at https://pypi.org/project/pyphetools/.

## Acknowledgments

Research reported in this publication was supported by the National Human Genome Research Institute (NHGRI) at the National Institutes of Health (NIH) under award nos. 1RM1HG010860 and 5U24HG011449 and by the National Institute of Child Health and Human Development (NICHD) at the NIH under award number 5R01HD103805.

